# Time warping between main epidemic time series in epidemiological surveillance

**DOI:** 10.1101/2023.02.07.23285605

**Authors:** Jean-David Morel, Jean-Michel Morel, Luis Alvarez

## Abstract

The most common reported epidemic time series in epidemiological surveillance are the daily or weekly incidence of new cases, the hospital admission count, the ICU admission count, and the death toll, which played such a prominent role in the struggle to monitor the Covid-19 pandemic. We show that pairs of such curves are related to each other by a generalized renewal equation depending on a smooth time varying delay and a smooth ratio generalizing the reproduction number. Such a functional relation is also explored for pairs of simultaneous curves measuring the same indicator in two neighboring countries. Given two such simultaneous time series, we develop, based on a signal processing approach, an efficient numerical method for computing their time varying delay and ratio curves, and we verify that its results are consistent. Indeed, they experimentally verify symmetry and transitivity requirements and we also show, using realistic simulated data, that the method faithfully recovers time delays and ratios. We discuss several real examples where the method seems to display interpretable time delays and ratios. The proposed method generalizes and unifies many recent related attempts to take advantage of the plurality of these health data across regions or countries and time, providing a better understanding of the relationship between them. An implementation of the method is publicly available at the *EpiInvert* CRAN package.

**Author summary:** To monitor an epidemic, it is crucial to understand the relationship between the incidence of new cases, the hospital admission count, the ICU admission count, and the death toll time series. The relationship between any pair of such indicators can be formulated in terms of temporal delays and ratios which evolve across time. Given two such time series, we develop, based on a signal processing approach, an efficient numerical method for computing their time varying delay and ratio curves. Using realistic simulated data, we show that the method faithfully recovers time delays and ratios. In addition, we discuss several applications to real epidemic data where the method seems to output interpretable time delays and ratios. The obtained relationship between these epidemic time series is a key issue in epidemiological surveillance. The proposed technique provides a new tool to visualize, compare, and understand the evolution of key epidemiological time series. An implementation of the method is publicly available at the *EpiInvert* CRAN package.

## Introduction

The COVID-19 epidemic has provided us with information, for many countries, on the evolution of a number of key instantaneous epidemic measurements such as the number of cases, deaths, hospitalizations, or intensive care units (ICUs) occupation. In this work, we explore the functional relationship between these time series. Let *f* (*t*) and *g*(*t*) be a pair of time series. We will assume that there exists a smooth time varying delay, *s*(*t*), between the values of both curves (such as the average temporal delay between cases and deaths). Moreover, we assume that once the delay between both curves is compensated, the ratio between their values, *r*(*t*), follows a smooth function that represents the causal relationship between both time series. Therefore we look for functions *s*(*t*) and *r*(*t*) such that

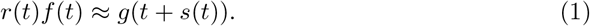

Of course, this problem is ill-posed because there are an infinite number of combinations of functions *r*(*t*) and *s*(*t*) that satisfy the above equation. Yet, one can transform the problem into a variational method where an energy is minimized. The first term of the energy is the quadratic difference between both members of (1) and the second term is a measure of the smoothness of functions *r*(*t*) and *s*(*t*), namely the quadratic norm of their derivatives. Minimizing jointly such an energy leads to find the smoothest time delays *s*(*t*) and smoothest ratio *r*(*t*) satisfying (1), by the classic Tikhonov-Arsenin solution for solving ill-posed problems [2]. The problem is then posed in terms of “we want the equation (1) to verify as well as possible but for *r*(*t*) and *s*(*t*) as smooth as possible”. In Figure 1 we illustrate an example of this approach when comparing, for the United Kingdom, between February 2020 and November 2023, the time series of cases and deaths. Key dates are indicated : the vertical bars indicate beginning or end of lockdown or social distancing measures and each shaded colored rectangle corresponds to a different wave, associated with the emergence of a different variant. We observe that the estimated *r*(*t*) and *s*(*t*) are quite smooth. A good agreement is obtained between *r*(*t*)*f* (*t*) and *g*(*t* + *s*(*t*)), except at the beginning of the epidemic where *r*(*t*)*f* (*t*) is much smaller than *g*(*t* + *s*(*t*)), and *s*(*t*) is negative. This fact is explainable by the very significant underestimation of the number of cases at the beginning of the epidemic, where the number of deaths was growing much faster than the capacity to test the population.

**Fig 1.**
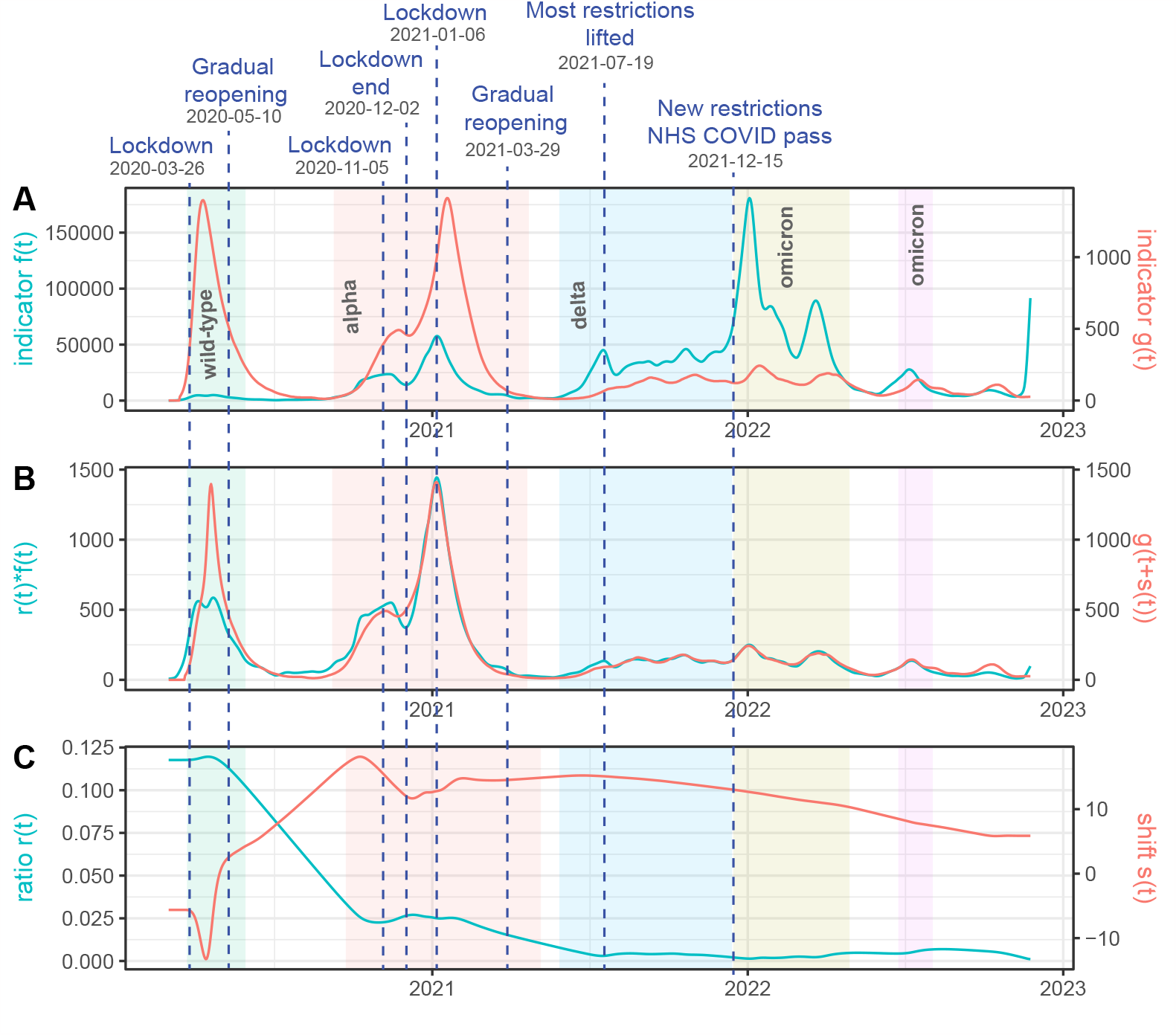
Illustration for the United Kingdom of the proposed warping method to compute the time warping *s*(*t*) and ratio *r*(*t*) relating the daily case incidence *f* (*t*) and the daily death count *g*(*t*) through the warping equation *r*(*t*)*f* (*t*) = *g*(*t* + *s*(*t*)). **(A)**: *f* (*t*) in cyan and *g*(*t*) in red. **(B)**: comparison of *r*(*t*)*f* (*t*) with *g*(*t* + *s*(*t*)) showing how well the warping equation is satisfied. **(C)**: the estimated *r*(*t*) in cyan and *s*(*t*) in red. Both are constrained by the method to be smooth to avoid overfitting. The color shaded rectangles represent the time intervals of the first five SARS-CoV-2 pandemic waves (as described in [1]).

Our proposed method aims at facilitating the comparison of epidemiological times series by computing and visualizing the latent variables *s*(*t*) and *r*(*t*) that link two time series. According to [3]:

Understanding and monitoring the epidemiological time delay dynamics of SARS-CoV-2 infection provides insights that are key to discerning changes in the phenotype of the virus, the demographics impacted, the efficacy of treatment, and the ability of the health service to manage large volumes of patients.

The authors of this paper studied how the pandemic has evolved in the United Kingdom through the temporal changes to the epidemiological time delay distributions between clinical outcomes. In [4] ten time periods were used to track changes in delays and the probabilities of outcomes using parametric statistical models in France. In each period, the variability of the delay was modeled using a parametric distribution. In [5], the difference between in-hospital mortality across four time periods was studied. [6] studied the evolution, in the USA, of in-hospital mortality by month of admission.

Comparison between epidemic waves is also a usual way to study the time evolution of epidemiological indicators. For instance, in [7], authors show that the in-hospital mortality was lower during the second wave of COVID-19 than in the first wave. Mortality predictors and differences in mortality between COVID-19 waves were identified using logistic regression in [8].

Despite these findings in specific examples, to our knowledge, no method exists to this date to continuously model and estimate the relationship between epidemiological curves. For example, the mortality rate may vary gradually depending on the improvement of medical treatments, the gradual introduction of vaccination or the gradual emergence of new variants. Until now, the way to study this variation has been to separate the epidemic time interval in different periods and assume a constant value in each period. This strategy requires selecting time periods, which is not an easy task because there exist many ways to do it, and calculating a constant value for each period. Knowledge of how the value of the variable varies within each period is lost in the process. In this paper, we propose a different approach where instead of separating the epidemic time interval in different periods, we model, using a generalized renewal equation, the evolution of the delay *s*(*t*) and the ratio *r*(*t*) between indicators as smooth functions defined in the whole epidemic time interval.

Our goal is to provide epidemiologists with tools providing a joint visualization, enabling the interpretation of two epidemiological curves related by causal relationship, such as incidence and death, or hospitalization and admission in ICU. While a direct visualization is possible through a mere superposition of two curves sharing the same time interval, we add and display in form of curves the two hidden variables connecting them : their time delay and their ratio. As we develop our Model section, Eq. (1) is an extension of the *renewal equation*, which is normally used for modeling a single incidence curve. This equation is extended here to relate two curves concerning the same population. In that way, we also extend and measure a generalized *reproduction number r*(*t*) which estimates (for example) an instantaneous ratio between incidence and mortality, and a generalized *time interval s*(*t*) that estimates for example the instantaneous time delay between detection and death.

## Model

### Dynamic time warping: the technique

Dynamic time warping (DTW) is a method to match two signals *f* (*t*) and *g*(*t*) by finding a nondecreasing shift function *w*(*t*) such that *f* (*t*) is as close as possible to *g*(*w*(*t*)). Variants of dynamic time warping like the one proposed in [9] match the slopes of the signals rather than the signals themselves. If the signals are sampled, thus encoded as two vectors, Bellman’s dynamic programming principle [10] applies and yields an optimal solution for any time discrete version of the cost *E*(*w*) =: *∫ c*(*f* (*t*) −*g*(*w*(*t*))^2^*dt*, where *c* is a positive cost function. As developed in [11], a discrete path optimization is performed in the 2D space of time correspondences, where only diagonal (*t, s*) → (*t* + 1, *s* + 1) horizontal (*t, s*) → (*t* + 1, *s*), and vertical (*t, s*) → (*t, s* + 1) moves are authorized. This discrete setting is efficiently solved by dynamic programming, and it can be extended to an optimal warping with time variable penalty [12]. We refer to [13] for a review of this algorithm. In this paper, we shall prefer a continuous formulation to the discrete one, in the line of [14], where a continuous time warping functional is defined, matching multiple copies of the same audio by a signal alignment that is robust to the presence of sparse outliers with arbitrary magnitude. The continuous formulations allow for a continuous iterative optimization by gradient descent and remove limitations on the form of the functional or discretization of the warp. As proposed in [15], a continuous penalty on the derivative of the warp can be used to ensure that this derivative does not vanish and does not become too high.

In the same line, the warping of signals has a 2D classic equivalent in the optical flow problem introduced in a seminal paper [16]. Applied to two successive images of a video, the method finds a vector field (2D warp) matching both images and enforces the warping field to be smooth by combining a regularity term with the matching fidelity term. In its initial formulation, the differential method worked only if the motion does not exceed one pixel. A more general computation of the optical flow is developed in [17] as a hierarchical multi-scale implementation of the optical flow using the Laplacian pyramid to represent both compared images at different scales. These ideas are developed in [18] which implements a multi-scale version of Horn-Schunk optical flow. The same hierarchical differential method is obviously applicable to pairs of similar signals.

### Epidemiological applications of time warping

In [19] the authors used dynamic warping to forecast the future spread of COVID-19 by exploiting the identified lead-lag effects between different countries. The past epidemic transmission delay between nations is determined through dynamic time warping between cumulative case curves. Then the cumulative case curve of the leading country is used to predict the Covid-19 spread in the following nation. In [20] the authors similarly present an algorithm for the clustering of confirmed COVID-19 cases at the county level in the United States. Dynamic time warping is used as a k-means clustering distance metric. The obtained clusters enable retrospective interpretation of the pandemic and informative inputs for case prediction models. In [21], the same method is developed for the Polish voivodeships. Similarities in time series are found by dynamic time warping between COVID-19 infections and deaths in pairs of vovoideships. The authors argue that a time warping method is important because the coronavirus epidemic did not start in all voivodeships at the same time. The method jointly analyzes the number of infected and deceased people in each province. Dynamic warping is applied to match pairs of (smoothed) incidence curves, pairs of cumulative case curves, and pairs of cumulative death curves.

In [22] the authors assessed the similarity between the time series of energy commodity prices and daily COVID-19 cases. They argue that the pandemic affects all aspects of the global economy and propose to assess the connections between the number of COVID-19 cases and the energy commodities sector. These connections are achieved by using Dynamic Time Warping to compute DTW distances between all time series—and use them to group the energy commodities according to their price change. In that way, the authors demonstrate that some commodities such as natural gas are strongly associated with the development of the pandemic. In [23] dynamic time warping was used to compare temporal patterns in patient disease trajectories. This enables the assessment of comorbidities in population-based studies, as it permits to identify more complex disease patterns than those derived from pairwise disease associations. The disease-history vectors of patients of a regional Spanish health dataset were represented as time sequences of ordered disease diagnoses. Statistically significant pairwise disease associations were identified and their temporal directionality was assessed. Subsequently, an unsupervised clustering algorithm, based on DTW, was applied on the common disease trajectories in order to group them according to their shared temporal patterns.

#### Limitations of time warping for epidemiological signals

We have listed in the preceding paragraphs the main computational contributions to time warping methods and several use cases in epidemiology. The main limitation of time warping comes from its equation. Given two signals *f* (*t*) and *g*(*t*), a (generally smooth) time shift *s*(*t*) is sought such that *f* (*t*) ≃ *g*(*t* + *s*(*t*)). This method is applicable to different but roughly proportional curves of the same phenomenon, for example to the incidence in neighboring regions. But it fails to fully model the relationship between curves that are linked to the same pandemic but are of different nature and scale, such as the pair formed by the incidence and death curves. While the examination of these curves suggests interpretable time delays, they also have a time varying relative rate, which may for example be linked to infectivity changes of a variant, to changes in the treatment, to vaccination, to evolution of the hospital treatment procedure, etc. Thus time warping, which shifts but preserves the values, is not applicable to such pairs. This is why we extend here the warping equation to introduce a varying rate or ratio *r*(*t*). The equation we aim at solving for each pair of curves is (1), namely

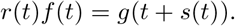

We shall relate this equation to the classic epidemiological renewal equation in the next paragraph. Yet, in the renewal equation the time interval is assumed known. Here *s*(*t*) could be interpreted as the mean of the time interval, but is unknown and must also be evaluated by the method. In the next paragraph we explain why (1) can be seen as a generalization of the renewal equation, and *r*(*t*) an extension of the reproduction number *R*_*t*_.

### The renewal equation

The reproduction number *r*(*t*) (in the literature often written *R*_*t*_ or *Rt*) is a key epidemiological parameter evaluating the transmission rate of a disease over time. It is defined as the average number of new infections caused by a single infected individual at time *t* in a partially susceptible population [24]. The reproduction number *r*(*t*) can be computed from the daily observation of the incidence curve *i*(*t*), but requires empirical knowledge of the probability distribution *k*(*s*) of the delay between two infections [25, 26].

There are two different models for the incidence curve and its corresponding infection delay *k*(*s*). In a theoretical model, *i*(*t*) would represent the real daily number of new infections, and *k*(*s*) is sometimes called *generation time* [27, 28] and represents the probability distribution of the time between infection of a primary case and infections in secondary cases, this time delay distribution being assumed stationary (independent of *t*). In practice, neither parameter is easily observable because the infected are rarely detected before the appearance of symptoms and tests will be negative until the virus has multiplied over several days. What is routinely recorded by health organizations is the number of *new detected, incident cases*. When dealing with this real incidence curve, *k*(*s*) is called *serial interval* [27, 28]. The serial interval is defined as the delay between the onset of symptoms in a primary case and the onset of symptoms in secondary cases [28].

A fundamental equation links *r*(*t*) to *i*(*t*) and *k*(*s*). It is the *renewal equation*, first formulated for birth-death processes in a 1907 note of Alfred Lotka [29]. We adopt the Nishiura et al. formulation [30, 31],

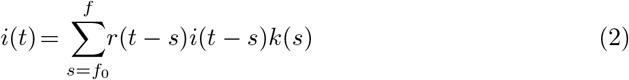

where *f*_0_ and *f* are the maximal and minimal observed times between a primary and a secondary case. In the renewal equation, the incidence curve *i*(*t*) is causally linked to itself through an internal delay represented by the probability distribution *k*(*s*) and a reproducing factor *r*(*t*). We now consider generalizations of this renewal that link time series that can still be related by causality, time delay, and a reproducing factor.

Our first example is the reproducing link from cases to deaths. Consider the time delay probability distribution *k*_*t*_(*s*) that a person reported dead at time *t* was reported positive at time *t* − *s*. Then we call *r*(*t*) the probability that a person diagnosed at time *t* will eventually die from the disease. With these assumptions, we obtain a *generation equation* linking cases to deaths, namely

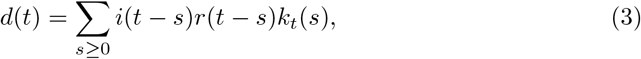

where *d*(*t*) denotes the daily death toll and *i*(*t*) the incidence curve. A strong simplification of the model may replace *k*_*t*_ by the Dirac mass concentrated at its center of mass *σ*(*t*) =: Σ _*s*_ *sk*_*t*_(*s*). Then (3) boils down to

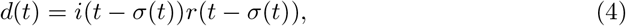

which amounts to assuming that all persons dead at time *t* have been detected at time *t* − *σ*(*t*). Let us now relate Eq. (4) to the simpler equation (1). This can be done by setting 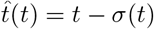. Then (4) rewrites

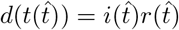

where 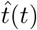 is the reverse change of variable. Setting 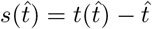 we get

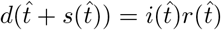

which is precisely (1). It remains to ask why the change of variable is licit. This is ensured if |*σ*′(*t*) | *<* 1, so that 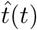 is strictly increasing. We shall ensure such a condition in our formalization of the problem.

### Mathematical formalization and resolution of (1)

#### The energy

Given two causally related time series *f* (*t*) and *g*(*t*), we introduced the problem of finding a smooth time warping *s*(*t*) and a smooth reproduction number *r*(*t*) such that

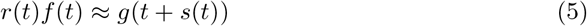

This leads us to solve the following variational formulation of the problem where *r*(*t*) and *s*(*t*) are estimated by minimizing the error functional

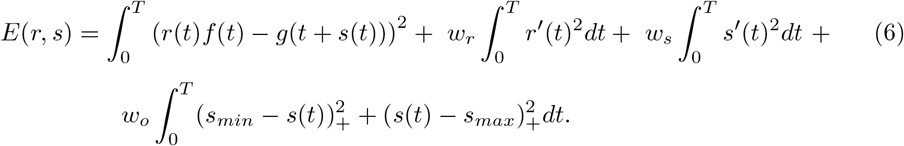

The first term of the energy (6) penalizes a poor matching between *r*(*t*)*f* (*t*) and *g*(*t* + *s*(*t*)). The second and third terms of the energy impose that *r*(*t*) and *s*(*t*) be smooth, and the strength of this smoothing is controlled by the weights *w*_*r*_ and *w*_*s*_. The larger the values of these weights the more regular *r*(*t*) and *s*(*t*) will be. The fourth term penalizes warps *s*(*t*) that would fall outside an expected interval [*s*_*min*_, *s*_*max*_] (the function (*x*)_+_ is defined as (*x*)_+_ = *x* if *x >* 0 and (*x*)_+_ = 0 otherwise).

#### Normalization and discretization

To compute the minima of the energy (6), we first normalize the time series *f* (*t*) and *g*(*t*) by dividing them by their medians. In this way, the minimization is independent of the time series magnitudes. Once the energy is minimized, the obtained value of *r*(*t*) is scaled to compensate for this normalization. Next, we express the energy in a discrete setting as

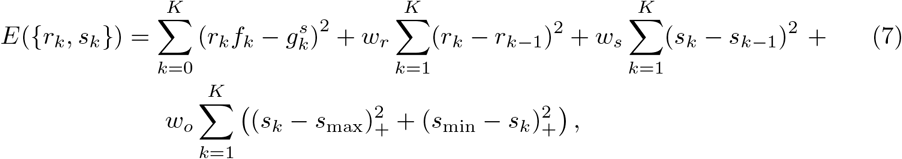

where *r*_*k*_ = *r*(*t*_*k*_), *s*_*k*_ = *s*(*t*_*k*_), *f*_*k*_ = *f* (*t*_*k*_), and 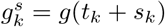.

#### Alternate minimization

We use an alternate iterative gradient descent type method to compute 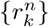 and 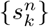. Given 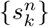 we compute 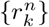 by expressing the derivatives of the energy with respect to 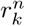 and equating them to 0. This yields a system of linear equations in 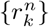,

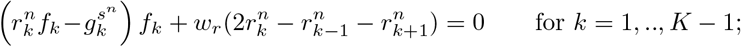

the cases *k* = 0 and *k* = *K* are managed using the boundary conditions. If the user has selected a value 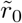 or 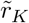 for *r*_0_ or *r*_*K*_, then we add to the system the associated equation 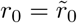 or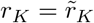. Otherwise we add the equations

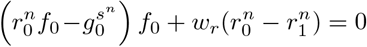

Or

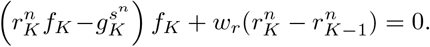

Given 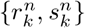, we compute 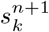 using a first order approximation of 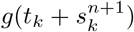. We express 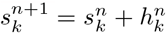 (then the unknown becomes 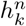) and we replace in the energy 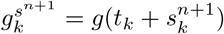 by 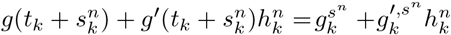, (where 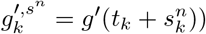). Differentiating the energy with respect to 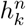 we obtain for *k* = 1, .., *K* − 1, a linear equation in 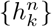,

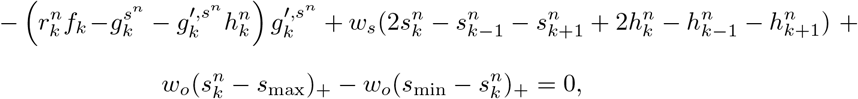

where the boundary conditions in *k* = 0 and *k* = *K* are managed in the same way as for 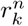. We perform iterations of the alternate scheme to compute 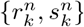 until convergence.

#### Initialization

Notice that we need to initialize 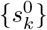 to start the iteration. We use two approaches to get an initialization of 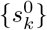. The first one is to put in correspondence the local maxima of *f* (*t*) and *g*(*t*). That provides the value of 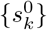 as the shift between the local maxima. For the rest of the points, we use linear interpolation to compute 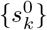 between the shift obtained for the local maxima. The second approach to initialize 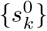 is to divide [0, *K*] in equally spaced subintervals [*k*_*m*_, *k*_*m*+1_] and for each subinterval, we compute 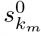 as the value *s* ∈ [*s*_min_, *s*_max_] which maximizes the covariance between 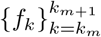 and 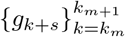. For the rest of points *k* ≠ *k*_*m*_ we initialize 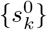 using linear interpolation. To get the best result for {*r*_*k*_, *s*_*k*_} we minimize the energy using both mentioned different strategies for the initialization of 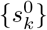 and we keep the one providing the lowest value of the energy.

#### Parameters

The main parameters of the energy are *w*_*r*_, *w*_*s*_, which determine the regularity of the ratio *r*(*t*) and of the time shift *s*(*t*), the interval [*s*_*min*_, *s*_*max*_] with determines the expected range of values for *s*(*t*), and the optional boundary values of *s*(*t*) and *r*(*t*) at the ends of the time interval [0, *T*]. All of these parameters can be tuned by the user. Unless otherwise indicated, the values of these parameters used in the experiments presented in this work are *w*_*r*_ = 1000, *w*_*s*_ = 10, [*s*_*min*_, *s*_*max*_] = [−10, 25] and, by default, optional boundary values are not used. The parameter *w*_*o*_ only intervenes in the case where *s*(*t*) goes out of the interval [*s*_*min*_, *s*_*max*_]. Since an interval [*s*_*min*_, *s*_*max*_], large enough to include the possible values of *s*(*t*) is normally used, the term in the energy with *w*_*o*_ is rarely used. We fixed *w*_*o*_ = 10^10^ in the energy minimization implementation.

## Material and methods

We can apply our method to any pair of epidemiological time series where we can expect that the generalized renewal equation (1) is satisfied. It is easily checked if the model provides a good matching between both time series (see Figure 1).

The time series to be compared must provide data with the same fixed time step (for instance, daily or weekly). Given the continuous formulation of the generalized renewal equation (1), as showed below in the experimental results, the method works better if the time series are corrected by adequate smoothing of the strong weekly oscillation, also called *administrative noise*.

All the experiments presented in this paper can be reproduced using the implementation of the method (named *EpiIndicators*) publicly available at the *EpiInvert* R CRAN package [32]. We used the values of the following time series provided by the Our World in data organization (**OWID**) [33] :

- *new_cases* : new daily confirmed cases of COVID-19
- *new_deaths* : new daily deaths attributed to COVID-19
- *icu_patients* : number of COVID-19 patients in intensive care units (ICUs) on a given day
- *hosp_patients* : number of COVID-19 patients in hospital on a given day
- *weekly_icu_admissions* : number of COVID-19 patients newly admitted to intensive care units (ICUs) in a given week (reporting date and the preceding 6 days)
- *weekly hosp admissions* : number of COVID-19 patients newly admitted to hospitals in a given week (reporting date and the preceding 6 days)

The raw registered number of daily cases and deaths are very irregular curves that contain a strong weekly seasonality due to the way countries register the number of cases and deaths every day of the week. To remove the noise and the weekly seasonality of these curves, we use, *EpiInvert*, the method proposed in [34] which estimates smooth trend curves for the incidence and death. In the experiments presented below, we use these restored versions of the incidence and deaths as the epidemic time series.

## Results

### Comparison of cases and deaths in the United Kingdom

In Figure 1 we show the results obtained when comparing cases and deaths in the United Kingdom. We shall compare these results with the ones obtained by other authors on related time series. In [3], the authors study the variation of the time delay between symptom onset and death in 4 epidemiological periods from 2020-01-01 to 2021-01-20 in the United Kingdom. In table 1 we present some of the results. Globally, the obtained time delay increases in a significant way in the first part of the year and declines in the last period. We compared this result with the one displayed in Figure 1. Note that the data are different: an incident case is recorded at the time of its registration while in [3] the case statistics is attributed to the time of symptom onset. We nevertheless observe the same trend: a significant increase in the first part of the year and a decline by the end of 2020. The maximum of our obtained delay, *s*(*t*), is 18.14 and the maximum of the time delay obtained in [3] is 24.69. This difference is justified by the delay between symptom onset and the inclusion of the case in the incidence statistics. This delay depends on the country’s data collection policy. Thus these data are too different to perform a quantitative numerical comparison of the results ; we limited ourselves to a study of the coherence of the results.

**Table 1.** Results, obtained in [3], for the mean of the time from symptom onset to death, segmented by symptom onset date using Lognornal distributions.

| Period | N. of samples | Mean |
| --- | --- | --- |
| 2020-01-01 to 2020-05-31 | 5378 | 19.61 |
| 2020-06-01 to 2020-08-31 | 170 | 24.69 |
| 2020-09-01 to 2020-11-30 | 1335 | 23.00 |
| 2020-12-01 to 2021-01-20 | 650 | 17.40 |

In [1], the authors estimate, for the United Kingdom, the death-rate, in the population, for the first five SARS-CoV-2 pandemic waves illustrated as color shaded areas in Figure 1. It should be noted that the article calculates the mortality rate of large population groups, without taking into account whether they have been infected or not. Therefore, the mortality rates are much lower than the ones computed in Figure 1, which refer to the incident population. Calculating the average of the *r*(*t*) calculated by our method in each period we obtain the comparison of Table 2. The results show good agreement, apart from two explainable discrepancies. In wave 1, the mortality rates diverge strongly because of the low testing capacity at the beginning of the epidemic. At this stage, most incident cases are only detected in hospitals while the results of [1] are based on a neutral population. In the later waves, the mortality rate obtained by [1] is about 10 times smaller than the one obtained on the incident population, with the exception of wave 4. This would mean that approximately 10% of the observed population was infected. In wave 4, the first wave of Omicron, more people were infected, which causes the rate given in [1] to go up, but the one measured on the incident population instead goes down, which is explainable by the fact that the first Omicron variant was very contagious but less lethal.

**Table 2.** Comparison, for the United Kingdom, of the death-rate per capita obtained in [1] for the first five SARS-CoV-2 pandemic waves and the average of *r*(*t*) in the same periods estimated using the case and deaths time series.

| Period | Variant | Death-rate [1] | Average $r(t)$ |
| --- | --- | --- | --- |
| 2020-03-23 to 2020-05-30 | wild-type | 0.00480 | 0.11502 |
| 2020-09-07 to 2021-04-24 | alpha | 0.00269 | 0.02371 |
| 2021-05-28 to 2021-12-14 | delta | 0.00064 | 0.00407 |
| 2021-12-15 to 2022-04-22 | omicron | 0.00101 | 0.00246 |
| 2022-06-22 to 2022-08-03 | omicron | 0.00067 | 0.00551 |

#### Comparison of hospitalizations and deaths

For France, we compare, in Figure 2, the hospitalizations and deaths restored using EpiInvert. In the data that we use, provided by the **OWID**, the hospitalizations are given by the weekly aggregated hospitalizations but updated every day with a different value, that is, we have a daily value of the time series. To get an approximation of the daily hospitalizations we divide this quantity by 7. In Figure 2 we observe a good agreement between *r*(*t*)*f* (*t*) and *g*(*t* + *s*(*t*)) with quite smooth *r*(*t*) and *s*(*t*).

**Fig 2.**
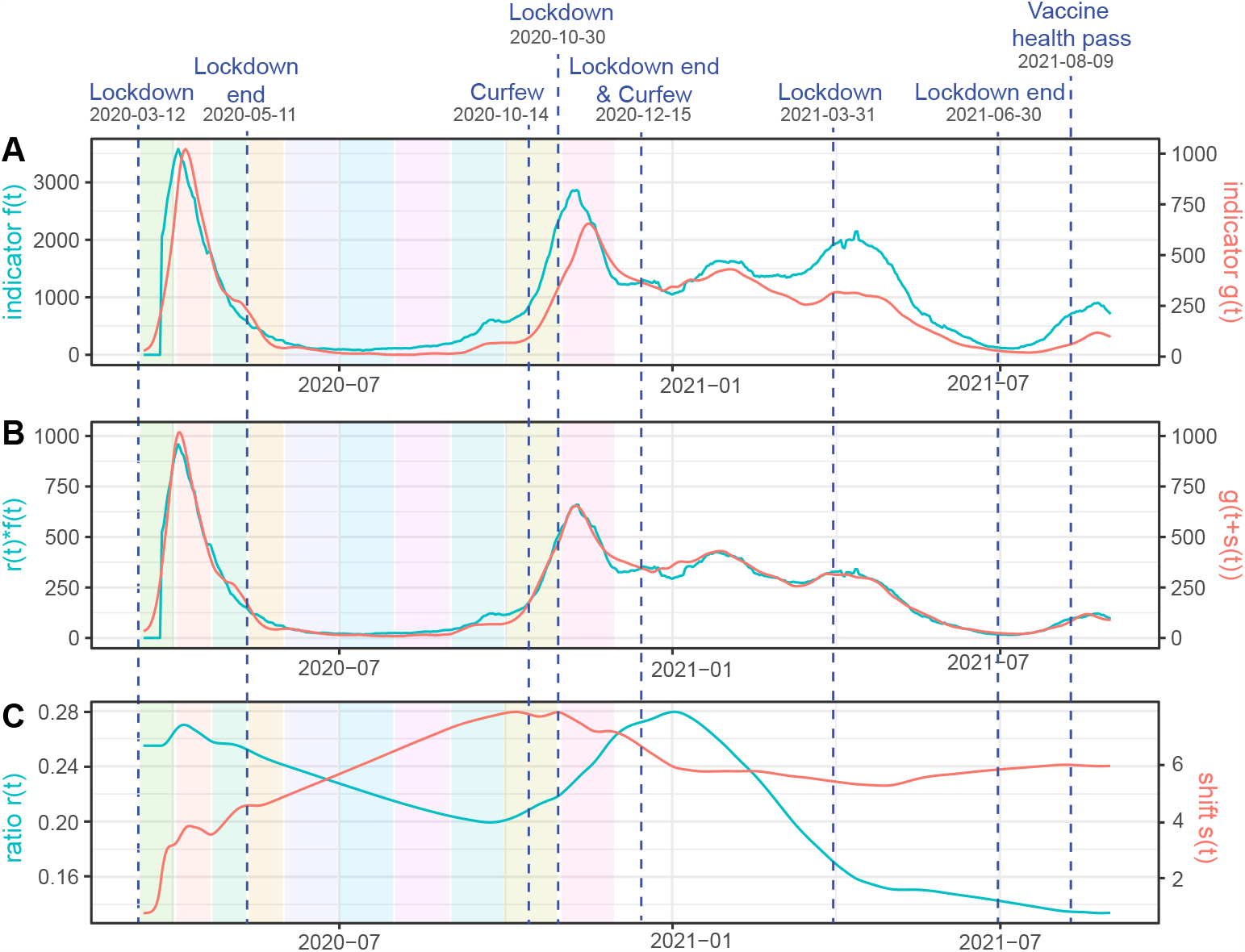
For France, we use as time series *f* (*t*) : the daily value of the weekly hospitalizations divided by 7 and *g*(*t*) : the restored number of daily deaths using EpiInvert. In **(A)** we plot *f* (*t*) and *g*(*t*). In **(B)** we plot the estimated *r*(*t*)*f* (*t*) and *g*(*t* + *s*(*t*)). In **(C)** we plot *r*(*t*) and *s*(*t*).

The colored rectangles in Figure 2 represent the time intervals used in [4] to study the variation of the mortality rate in hospitals. in Figure 3 we present a comparison between the mortality rate obtained in [4] and the value of *r*(*t*) showed in Figure 2. Notice that the data are quite different because we use the registered values of weekly hospitalization and deaths and the authors of [4] use the values obtained on a cohort of patients. The reporting delays of the data can be also quite different for each kind of data. With this caveat, the obtained comparative results show a similar order of magnitude. Taking into account that the periods used in [4] are very short (the first six periods belong to the first wave), a mortality decay from 25% to 10% in a few months seems excessive. In the first period for example, in less than 3 weeks and at the beginning of the epidemic, mortality goes down from 24.9% to 20.6%, which is not very credible, especially in the first weeks when everything was getting worse. In that sense, our result that shows a worsening death rate at the beginning before starting to decrease seems more reasonable. We can add to the preceding caveat that the number of cases is very small between sections 5 and 7, which deteriorates the quality of the results when calculating separately by sections. The trend is nevertheless similar in both estimates: for the first variant, the mortality decreases progressively during the first wave, to grow again from the emergence of the Alpha variant in the fall of 2020. At the peaks of the waves, the results of both estimates are closer to each other, which is logical as more data were available.

**Fig 3.**
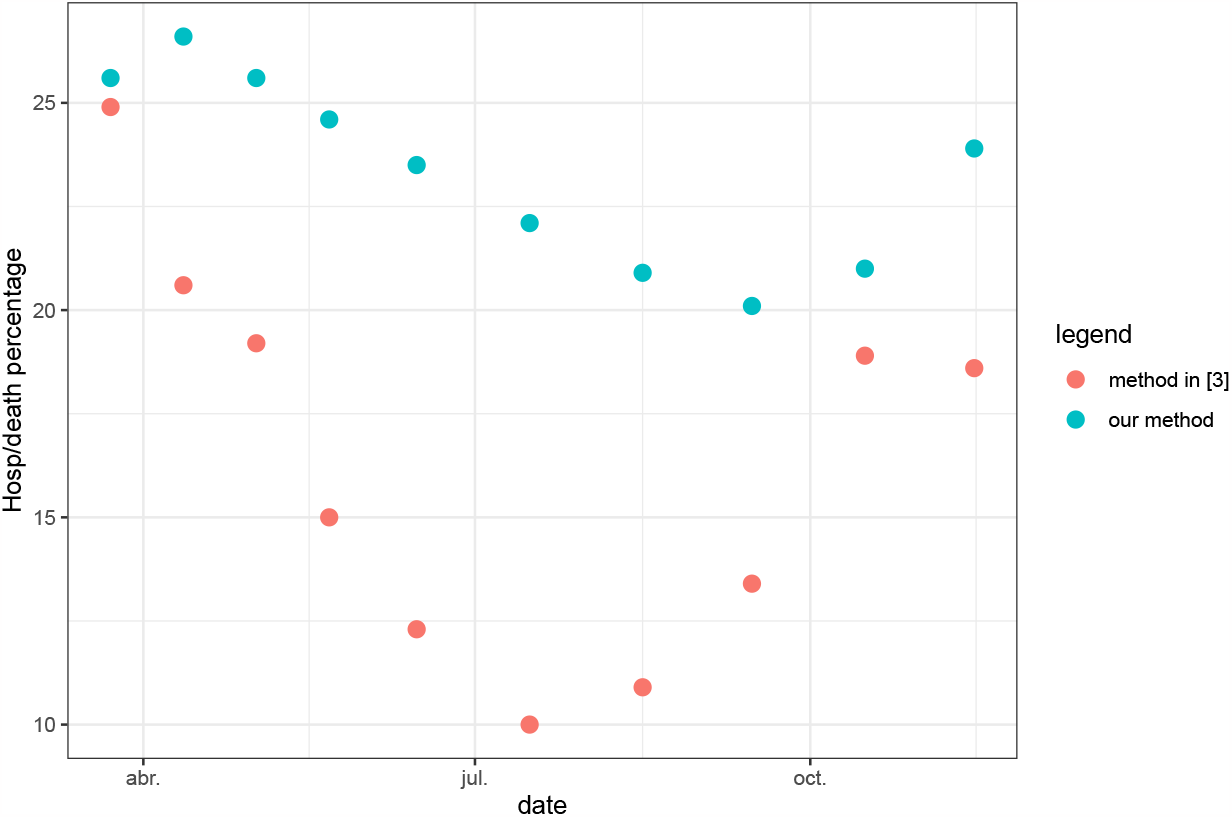
For France, we show the evolution, during the first epidemic year of the mortality rate in hospital obtained in [4] and the average of *r*(*t*) in the same periods showed in Figure 2

#### Influence of the weights *w*_*r*_ and *w*_*s*_

The lower the values of the weights *w*_*r*_ and *w*_*s*_, the better the match between *f* (*t*)*r*(*t*) and *g*(*t* + *s*(*t*)), but the functions *r*(*t*) and *s*(*t*) are more irregular. At the end of the paper we include a justification of the default choices of these parameters using realistic simulated data. To illustrate the influence of such choices, we compare in Figure 4 hospitalizations and deaths in France when lowering *w*_*r*_ and *w*_*s*_. As expected, the match between *r*(*t*)*f* (*t*) and *g*(*t* + *s*(*t*)) is better than in Figure 2, but *r*(*t*) and *s*(*t*) are more irregular. In fact, we observe significant oscillations of *r*(*t*) and *s*(*t*) in very short periods of time that suggest that the values for *w*_*r*_ and *w*_*s*_ are too low in this experiment, so that both corrected curves overfit.

**Fig 4.**
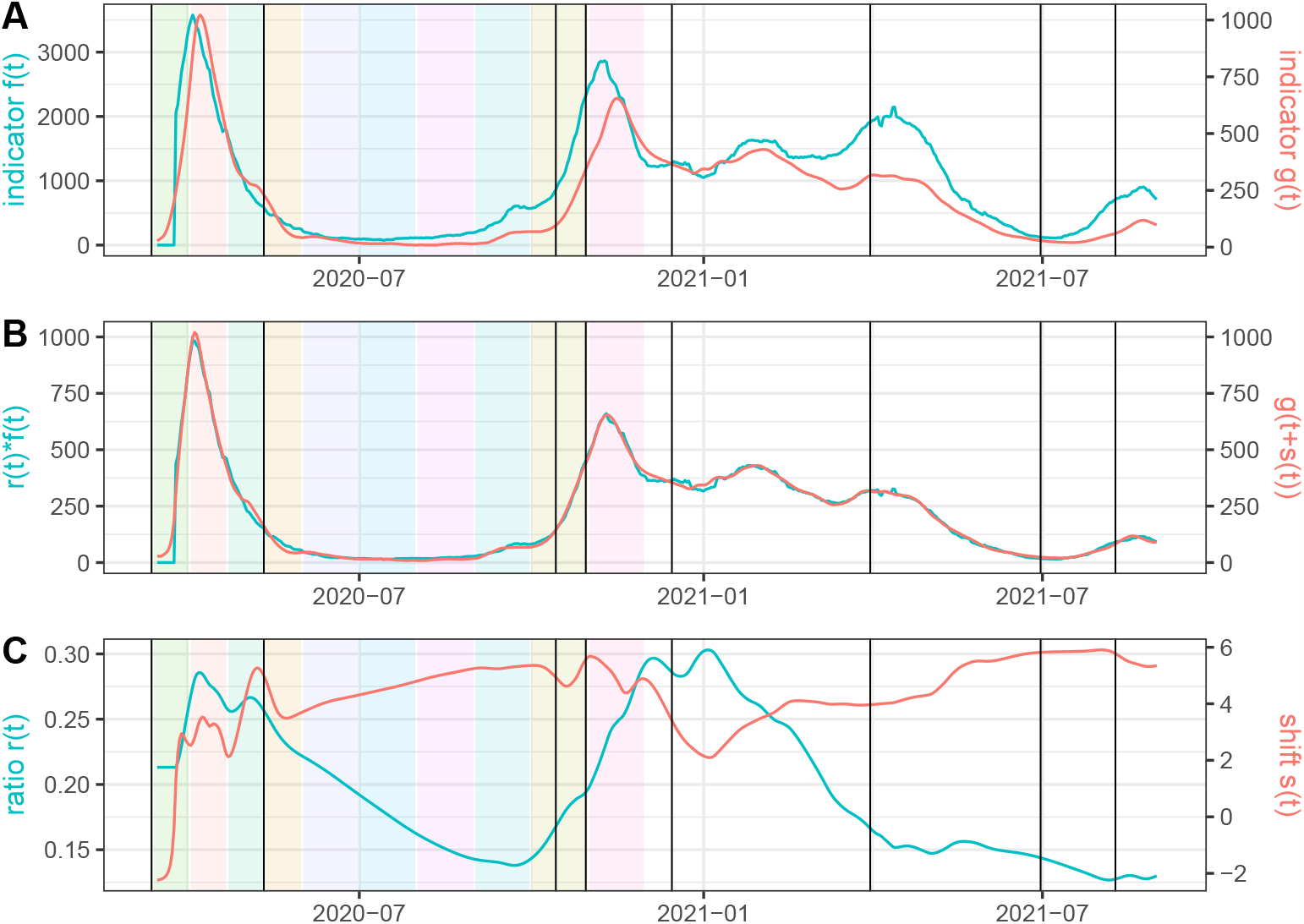
Same experiment as in Figure 2 but using *w*_*r*_ = 10 (instead of the default value *w*_*r*_ = 1000) and *w*_*s*_ = 1 (instead of the default value *w*_*s*_ = 10).

### Comparing the result of *EpiInvert* with the smooth values of cases provided by the OWID

As stated above, the regularization of the raw incidence and death data is an important pre-processing step to improve the quality of the results. In this experiment, we compare the influence of the regularization step when processing the same raw time series with two different methods. In Figure 5 we compare the smooth version of the raw incidence provided by the **OWID** in France with the smooth version of the raw incidence obtained by EpiInvert, a functionality included in the EpiInvert CRAN package [32]. We observe that the **OWID** smooth version of cases is more irregular than *EpiInvert*, it can be zero sometimes and the median of the delay *s*(*t*) is about 1.81 days. This experiment also illustrates another use of our method: comparing different versions of a same epidemiological curve and quantifying their difference.

**Fig 5.**
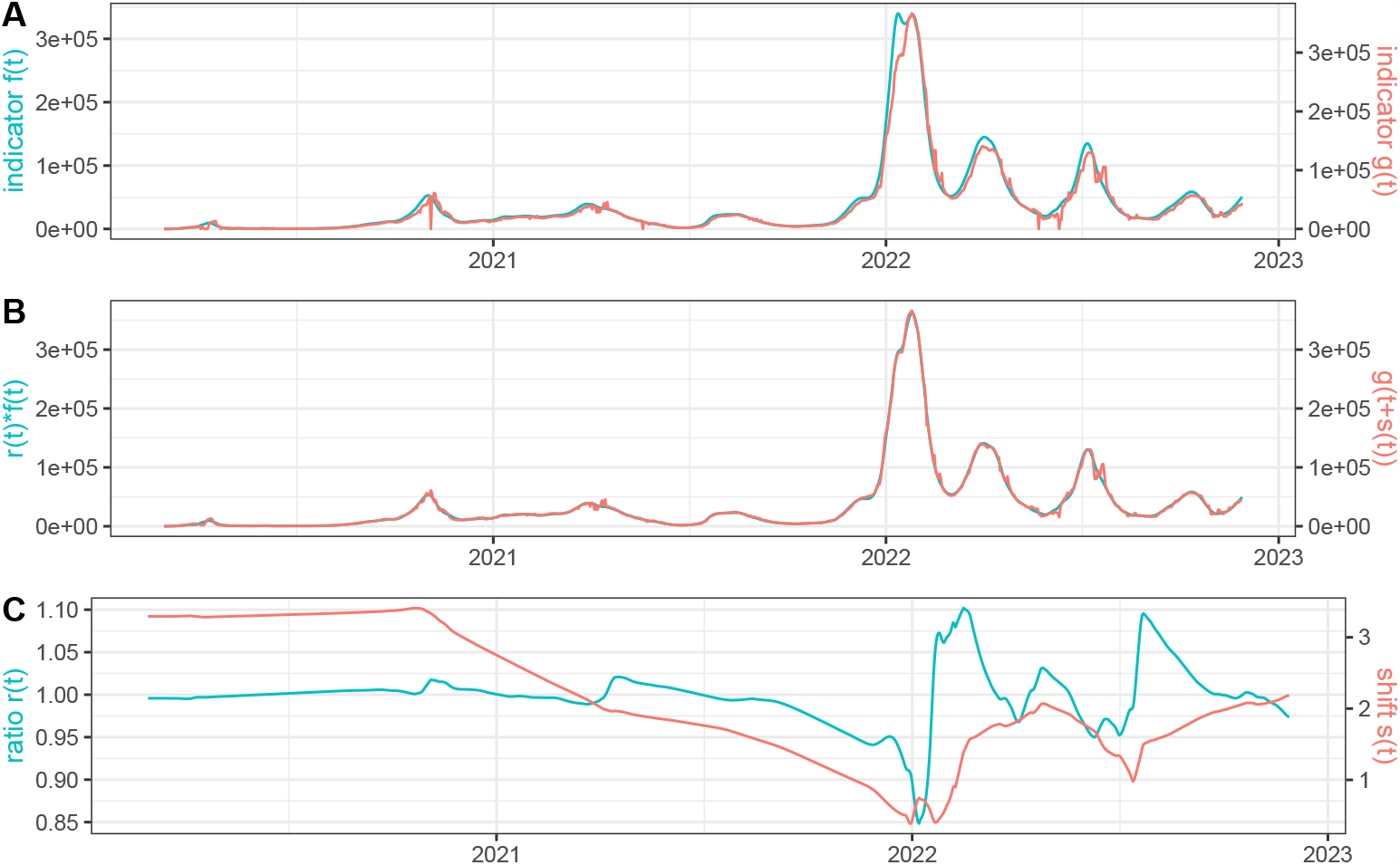
We use as time series *f* (*t*): the restored number of daily cases in France using EpiInvert and *g*(*t*): the smooth values of daily cases provided by the **OWID**. In **(A)** we plot *f* (*t*) and *g*(*t*). In **(B)** we plot the estimated *r*(*t*)*f* (*t*) and *g*(*t* + *s*(*t*)). In **(C)** we plot *r*(*t*) and *s*(*t*).

#### Verifying the model by a simulation study

To evaluate the capacity of *EpiIndicators* to correctly estimate *r*(*t*) and *s*(*t*) we developed a realistic simulation framework where both functions were known. We used as second time serie *g*(*t*) the restored number of deaths in France using *EpiInvert* and we generated the first time serie *f* (*t*) from simulated *r*(*t*) and *s*(*t*) using the formula

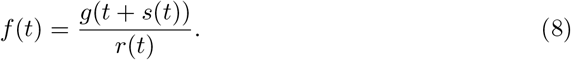

in this way, we obtained a perfect matching between both curves. The function *r*(*t*) was obtained using ‘atan()’ type functions where we simulated a decreasing behavior for *r*(*t*), *s*(*t*) was simulated in the same way with an increasing behavior. After simulating *f* (*t*) and *g*(*t*) in that way with a “ground truth” (*s*(*t*), *r*(*t*)), we applied *EpiIndicators* to the pair *f* (*t*), *g*(*t*) and compared the obtained values of *r*(*t*) and *s*(*t*) with the ground truth. The results are shown in Figures 6 and 7. We observe a good agreement between the estimated values of *r*(*t*), *s*(*t*), and the ground truth. Notice that we cannot expect the values to be exactly equal, because in the areas where *g*(*t* + *s*(*t*)) is very small or constant, there is not enough information to compute *s*(*t*). In that case, the energy regularity term dominates, causing the estimated value to be slightly smoother than the ground truth. On the contrary, at the peaks of the epidemic waves, the information is more robust and we can expect the calculation of *r*(*t*) and *s*(*t*) to be more accurate.

**Fig 6.**
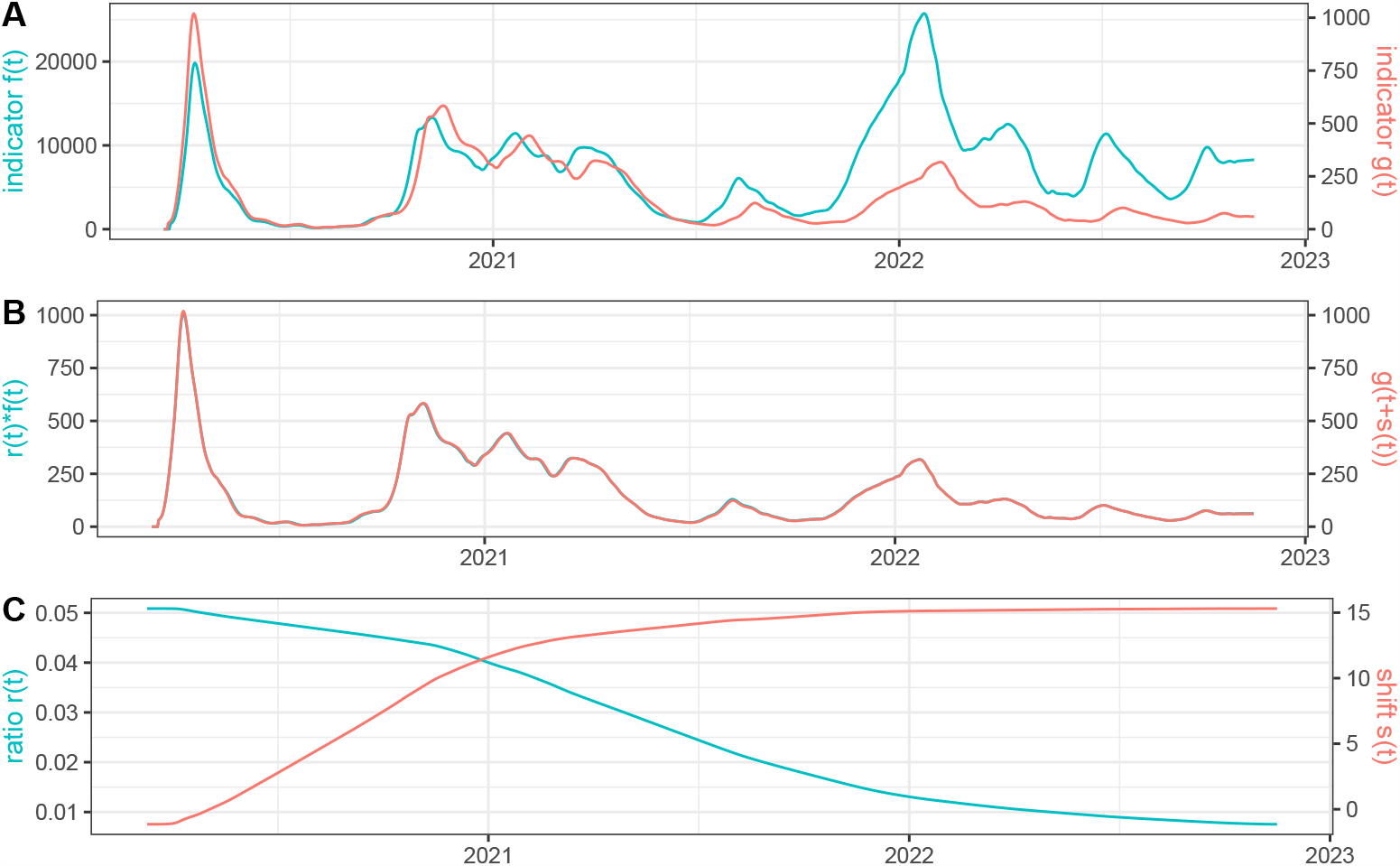
We use as time series *g*(*t*) the restored number of deaths in France using *EpiInvert* and *f* (*t*) is defined as 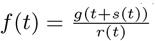 where *s*(*t*) and *r*(*t*) are a simulated ground truth. In **(A)** we plot *f* (*t*) and *g*(*t*). In **(B)** we plot the estimated *r*(*t*)*f* (*t*) and *g*(*t* + *s*(*t*)) using the recovered values for *r*(*t*) and *s*(*t*). In **(C)** we plot the recovered values for *r*(*t*) and *s*(*t*).

**Fig 7.**
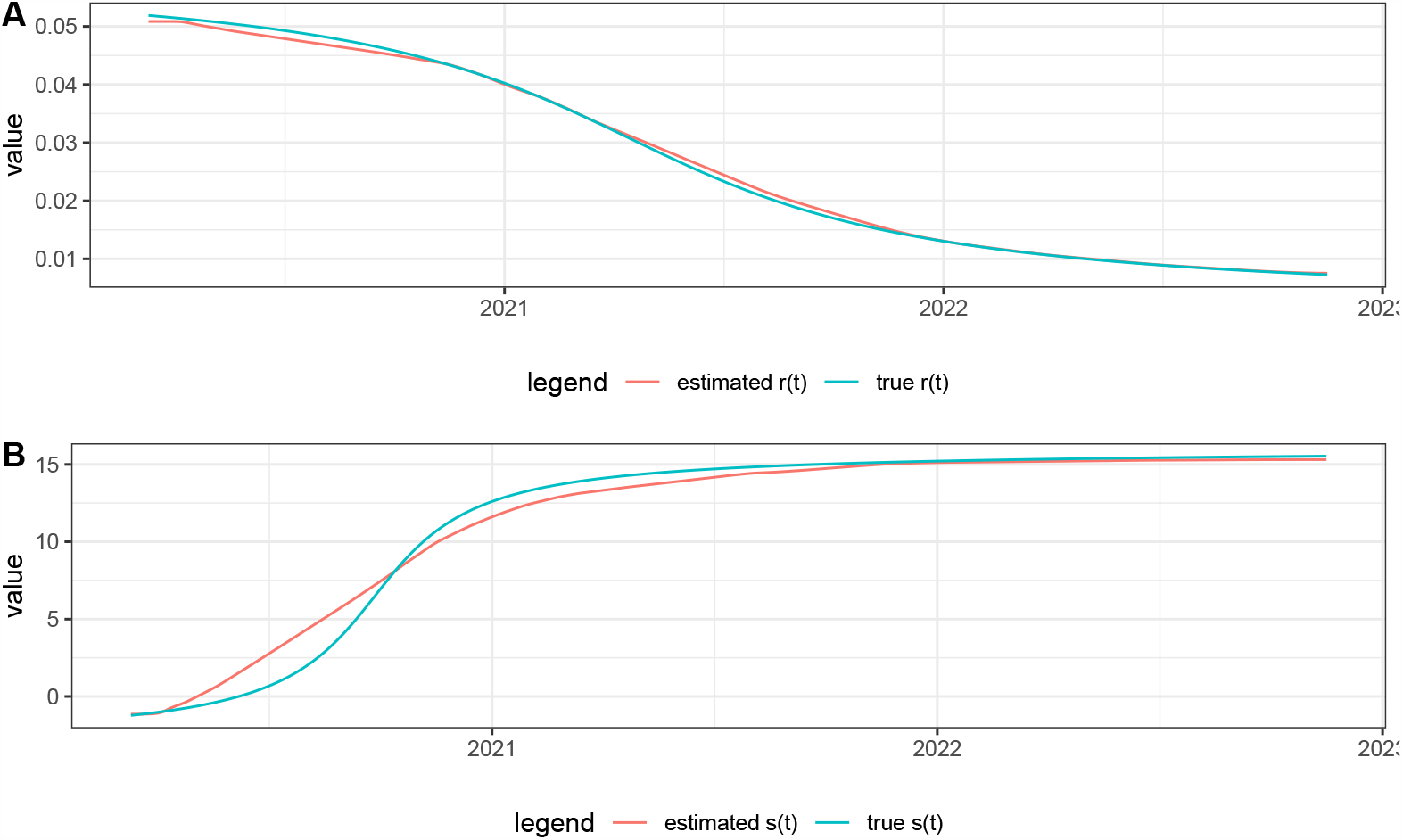
Comparison of the values of *r*(*t*) and *s*(*t*) estimated by the model and their ground truth using the simulated data.

To illustrate the influence of the regularization of the time series in the quality of the *r*(*t*) and *s*(*t*) estimations, we show in Figure 8 the results obtained using the same simulation but using as function *g*(*t*) the original raw number of reported deaths instead of the regularized version using *EpiInvert*. We observe that the estimations of *r*(*t*) and *s*(*t*) are strongly perturbed by the noise and weekly bias of the raw data: compare with the results in Figure 6 and 7.

**Fig 8.**
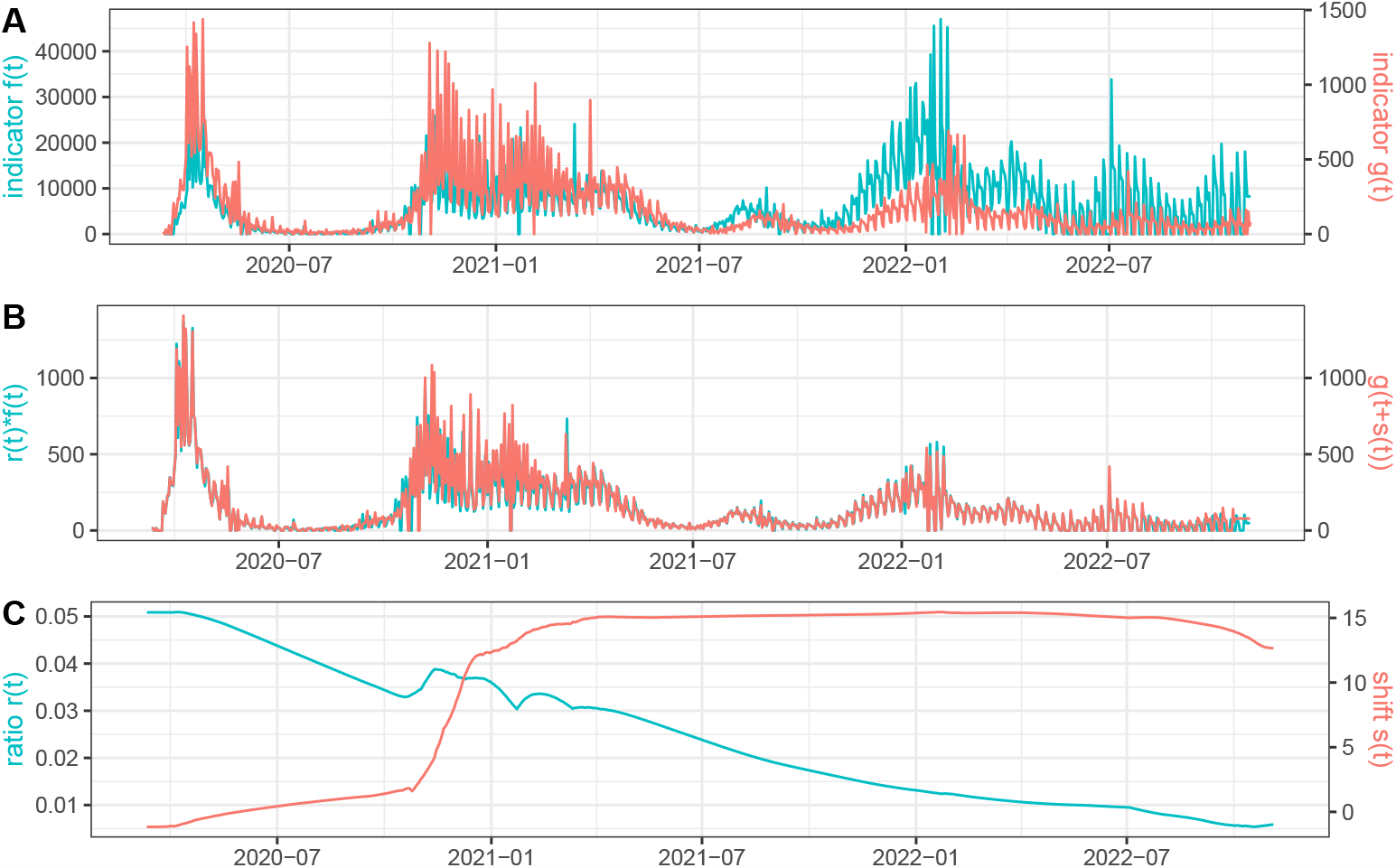
We use as time series *g*(*t*) the raw number of reported deaths in France and *f* (*t*) is defined as 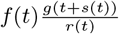 where *s*(*t*) and *r*(*t*) are a simulated ground truth. In **(A)** we plot *f* (*t*) and *g*(*t*). In **(B)** we plot the estimated *r*(*t*)*f* (*t*) and *g*(*t* + *s*(*t*)) using the recovered values for *r*(*t*) and *s*(*t*). In **(C)** we plot the recovered values for *r*(*t*) and *s*(*t*).

Next, we studied, using these simulated data, the influence of the order in which the time series are taken. Let 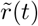 and 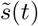 be the ratio and delay obtained using *g*(*t*) as the first curve and *f* (*t*) as the second curve. Then, accordingly with our model, we have that

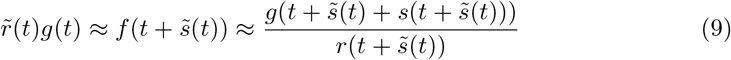

To check if this relation is satisfied we studied the normalized error distribution

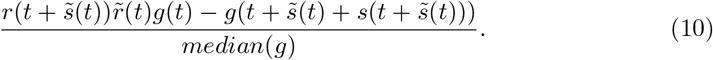

The results are shown in Figure 9. The small size of the normalized error (10) shows the performance of the method and its stability when we change the role of the time series. In the beginning, we find larger errors due to boundary effects in the estimations.

**Fig 9.**
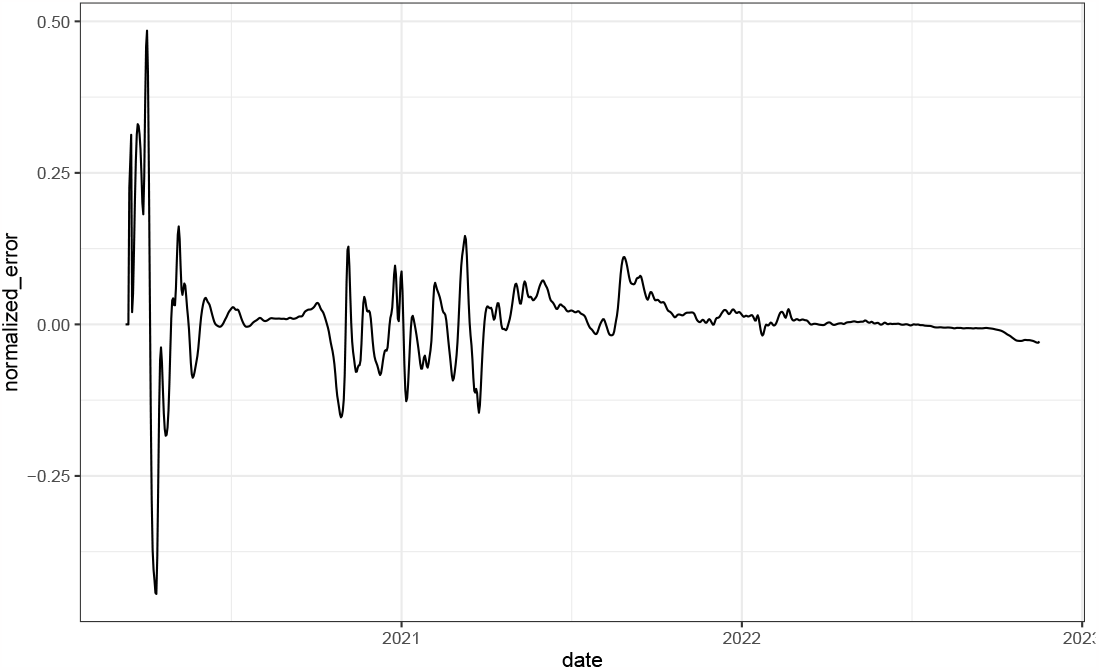
Normalized error (10) using the simulated data.

### A sanity check: verifying the transitivity of the estimations of *r*(*t*) and *s*(*t*)

In this paragraph, we study the transitivity of the estimation when three time series are involved. Let *f*_1_(*t*), *f*_2_(*t*) and *f*_3_(*t*) be these three time series. We took as example the France incidence curve *f*_1_(*t*) restored using *EpiInvert* ; *f*_2_(*t*) is the number of new hospital admissions and *f*_3_(*t*) is the number of new deaths restored using *EpiInvert*. The hospitalization data was taken from the COVID-19 European Hub. We will study the relationship between the ratio and delay between the three time series. Let *r*_*i,j*_(*t*) and *s*_*i,j*_(*t*) be the ratio and delay between time series *f*_*i*_(*t*) and *f*_*j*_(*t*). Then we have

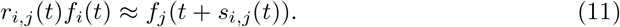

Using this relation for {*i, j*} = {1, 2}, {2, 3}, {1, 3} we obtain the following transitivity relation:

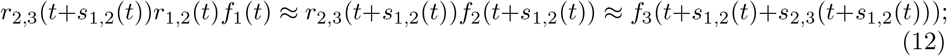

therefore the transitivity condition can be expressed using the equations

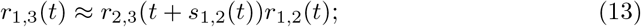

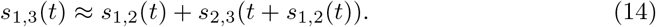

Next, we check if the relations (13) and (14) were satisfied for the proposed example. To do so we estimated *r*_*i,j*_(*t*) and *s*_*i,j*_(*t*) using the proposed method. Then, in Figure 10, we plotted *r*_1,3_(*t*) and *r*_2,3_(*t* + *s*_1,2_(*t*))*r*_1,2_(*t*) in (**A**) and *s*_1,3_(*t*) and *s*_1,2_(*t*) + *s*_2,3_(*t* + *s*_1,2_(*t*)) in (**B**). We notice a reasonable transitivity between the estimations of the ratio and delay for the three time series, except at the beginning of the epidemic, likely due to the poor quality of the initial incidence curve.

**Fig 10.**
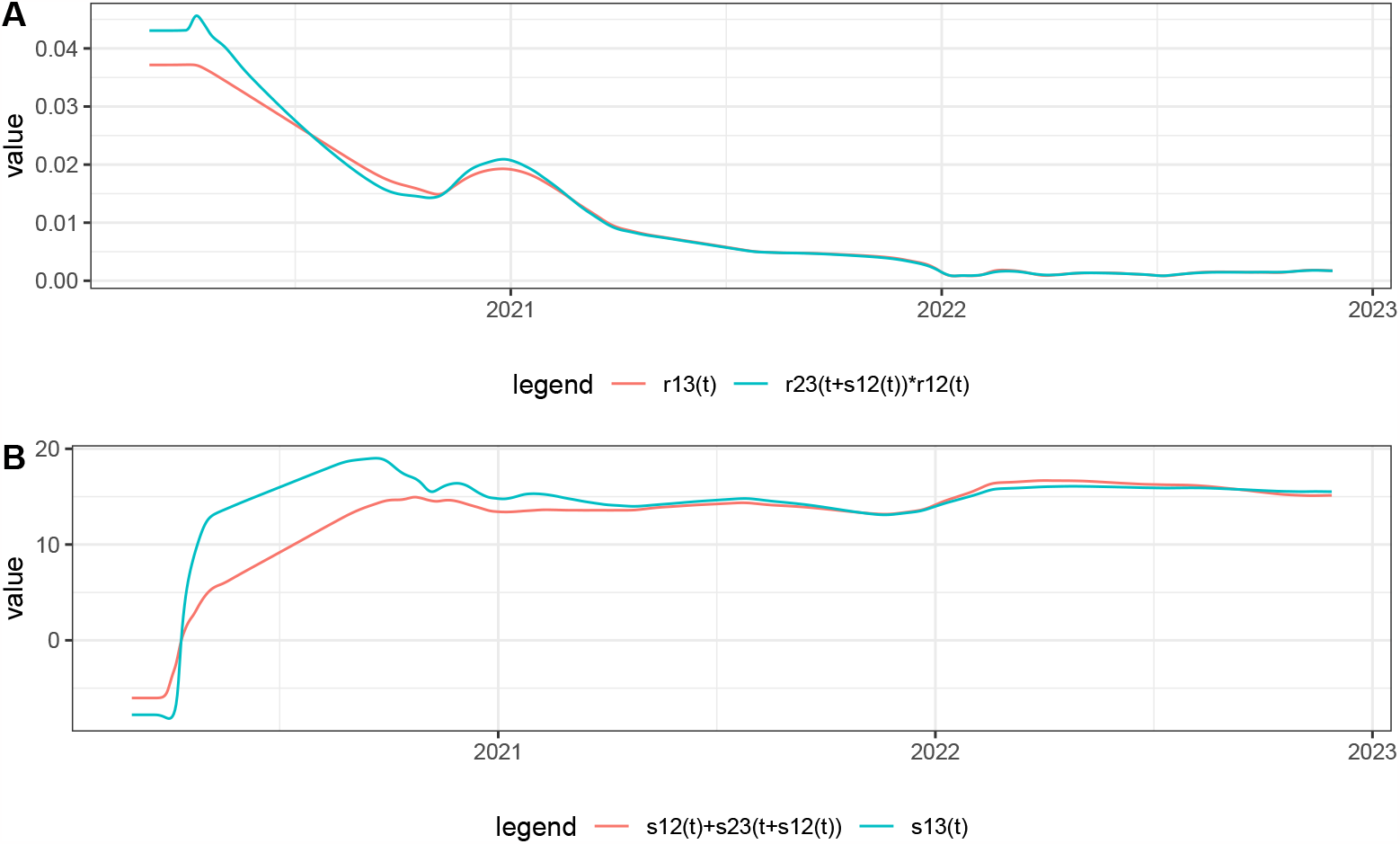
A sanity check: verifying the transitivity of the estimations of *r*(*t*) and *s*(*t*) when three time series are involved: *f*_1_(*t*), the France incidence curve, *f*_2_(*t*), the number of new hospital admissions and *f*_3_(*t*) the number of new deaths. In (**A**) we plot *r*_1,3_(*t*) and *r*_2,3_(*t* + *s*_1,2_(*t*))*r*_1,2_(*t*) and in (**B**) we plot *s*_1,3_(*t*) and *s*_1,2_(*t*) + *s*_2,3_(*t* + *s*_1,2_(*t*)).

### Comparison of epidemiological waves between countries

In Figure 11, we use our method to compare the deaths between Italy and France. It must be pointed out that this comparison is not fully legitimate. The populations being distinct, there is no formal argument to involve a generalized renewal equation between both incidence curves. Furthermore, the health policies in France in Italy were different. This application of our method is justified by the fact that we can expect a similar trend of the epidemiological time series in neighboring countries modulus a shift given by the temporal difference in the entry of the different variants of the virus in the countries. Variations in *r*(*t*) can be expected due to the different health policies. Indeed, the results in Figure 11 show a very good agreement between *r*(*t*)*f* (*t*) and *g*(*t* + *s*(*t*)), *s*(*t*) is quite smooth and *r*(*t*) is quite stable, oscillating most of the times in a short range of values ([0.8, 1.2]). The epidemic started in Italy before France. This is reflected in that *s*(0) ≈ 11.8 days. Then *s*(*t*) decreases sharply and stabilizes near zero, indicating a coordination of the epidemic waves in both countries. We also know that the first wave of the epidemic in Italy generated a large number of deaths; this is reflected in that *r*(0) ≈ 1.12. The median of *r*(*t*) for the whole period is 0.885, which is very close to the ratio between the populations of Italy and France (around 0.875), which suggests that, globally, the mortality rate in both countries has been similar. The factor 1.2 in the first three months of the pandemic indicates a larger mortality in Italy where only adaptive regional lockdown was applied. In France a strict lockdown was applied in the corresponding period. We can conclude that the overall mortality in France and Italy was proportional to their population, in spite of different policies.

**Fig 11.**
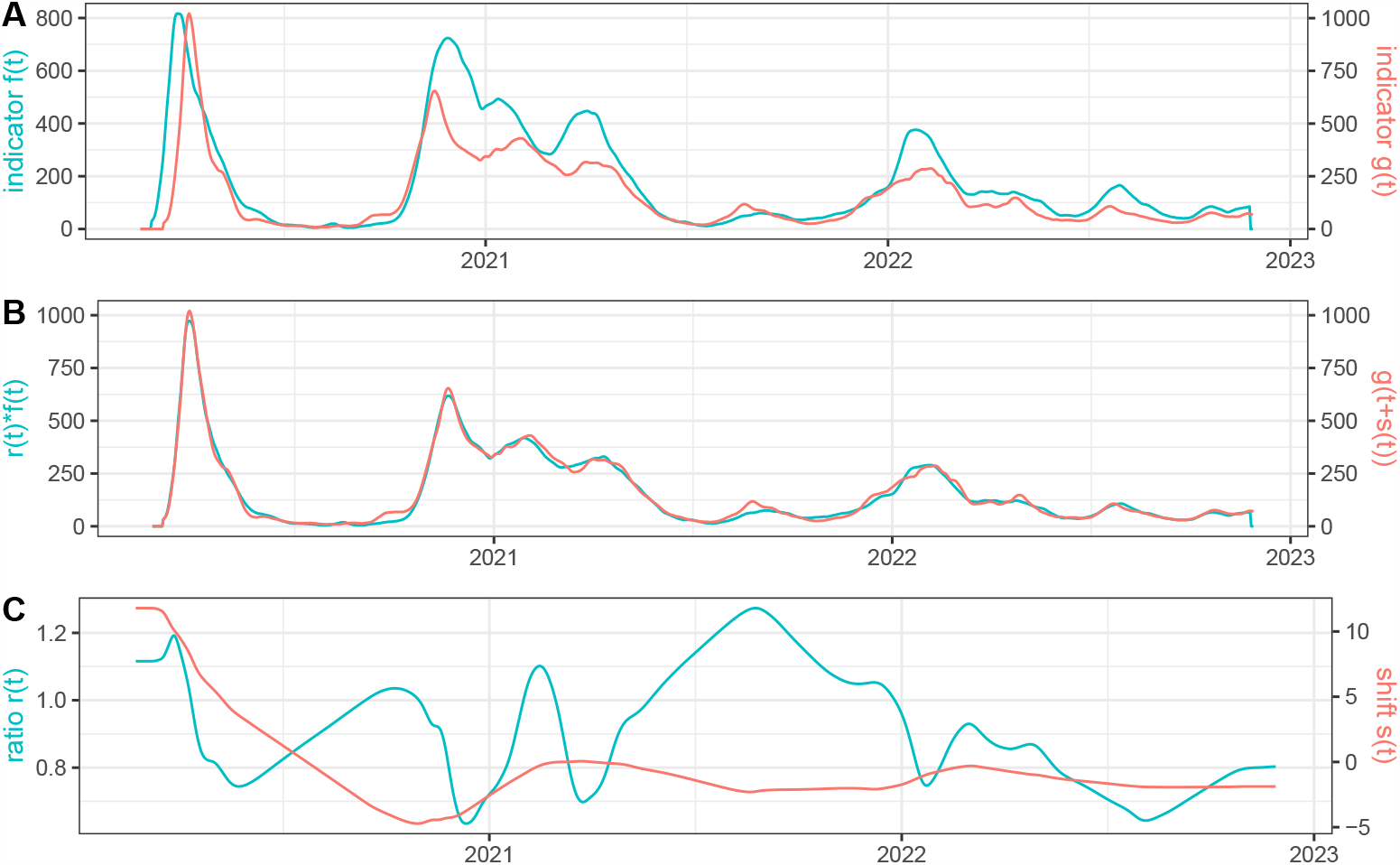
We use as time series *f* (*t*) : the restored number of daily deaths in Italy using EpiInvert and *g*(*t*) : the restored number of daily deaths in France using EpiInvert. In **(A)** we plot *f* (*t*) and *g*(*t*). In **(B)** we plot the estimated *r*(*t*)*f* (*t*) and *g*(*t* + *s*(*t*)). In **(C)** we plot *r*(*t*) and *s*(*t*).

### Applications to other time series

It might be argued that the generalized renewal equation proposed here, *r*(*t*)*f* (*t*) = *g*(*t* + *s*(*t*)) might be applied to more general situations than pandemic-related curves, namely any case where two curves are causally related, such as temperature and flu incidence. We refer to [35] for a study of the climate influence on dengue propagation in Peru. This study estimated the timing difference of dengue epidemics between jungle and coastal regions, with differences significantly associated with the seasonal cycle of mean temperature. We took a similar but more straightforward example and performed a comparative analysis of the weekly France flu incidence (data obtained at *sentiweb*.*fr*) and the corresponding weekly average temperature evolution (data obtained at *opendatasoft*.*com*) in the 2010-2019 period. Figure 12 (A) displays the cyan incidence curve and a red oscillatory curve corresponding to *f* (*t*) = *max*(*T* (*t*)) − *T* (*t*) where *T* (*t*) is the temperature. This inversion ensures that low temperatures correspond to high values of *f* (*t*) and enables a direct application of our proposed warping method ensuring that *r*(*t*) is positive. A visual examination of both curves shows a coincidence between yearly temperature minima and flu incidence peaks with a short delay. Figure 12 (B) displays a comparison of both curves after applying the estimated temporal shift *s*(*t*) and ratio *r*(*t*). This result nevertheless shows an imperfect match. This fact, in addition to the high oscillatory behavior of the red curve *s*(*t*) and the cyan curve *r*(*t*) in Figure 12 (C) means that our model does not provide a good result matching temperature and flu incidence. Indeed, we find that, in general, the temperature is not proportional to the flu incidence modulus a shift *s*(*t*). The ratio *r*(*t*) and the time delay *s*(*t*) provide no more meaningful insight than what was known beforehand, that low temperatures favor the emergence of flu. This negative experiment leads us to restrict the application of our method to cases where the compared curves are legitimately linked by a general renewal equation of the proposed form, *r*(*t*)*f* (*t*) = *g*(*t* + *s*(*t*)).

**Fig 12.**
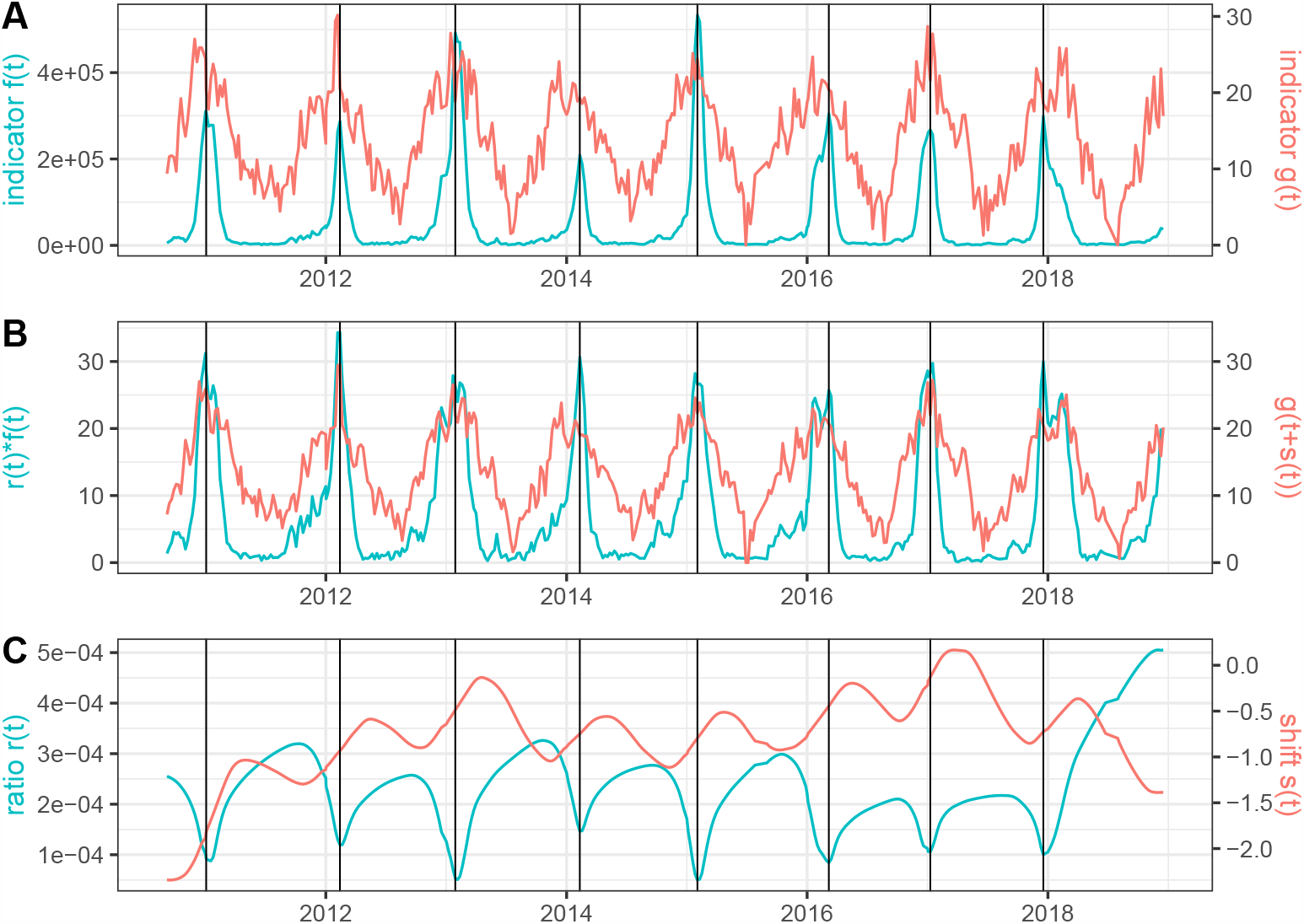
Results obtained using as *f* (*t*) the weekly flu incidence and *g*(*t*) = *max*_*t*_(*T* (*t*)) *− T* (*t*) where *T* (*t*) is the weekly average of the temperature in France. The black vertical lines correspond to the peaks of the flu incidence.

### Justification of the choice of the regularization weights *w*_*r*_ and *w*_*s*_

To justify the choice of the regularization weights *w*_*r*_ and *w*_*s*_ we shall use the realistic simulated data shown in Figure 6. The advantage of this simulated data is that we know the ground truth for *r*(*t*) and *s*(*t*). We can therefore compute the *RMSE* between this ground truth and the method’s *r*(*t*) and *s*(*t*) estimations for arbitrary choices of *w*_*r*_ and *w*_*s*_. To study the influence of *w*_*r*_ and *w*_*s*_ on the *RMSE* we added a perturbation to *f* (*t*) and *g*(*t*) given by a multiplicative Gaussian noise with mean 1 and standard deviation 0.3. On the results, we also applied a 7-day centered window average. To get an intuitive relative error for *r*(*t*), we divided the *RMSE* by the mean of the true *r*(*t*). For *s*(*t*) this normalization step was not needed because the reference time unit is always the day. in Figure 13 and 14 we show the obtained *RMSE*s. We find a number of combinations with accurate results using *w*_*r*_ between 100 and 10000 and *w*_*s*_ between 1 and 100. The results are therefore quite stable in a broad range of values of *w*_*r*_ and *w*_*s*_. In particular, the default choices of the parameters *w*_*r*_ = 1000 and *w*_*s*_ = 10 provide the best *RMSE* result for *r*(*t*) and among the best ones for *s*(*t*).

**Fig 13.**
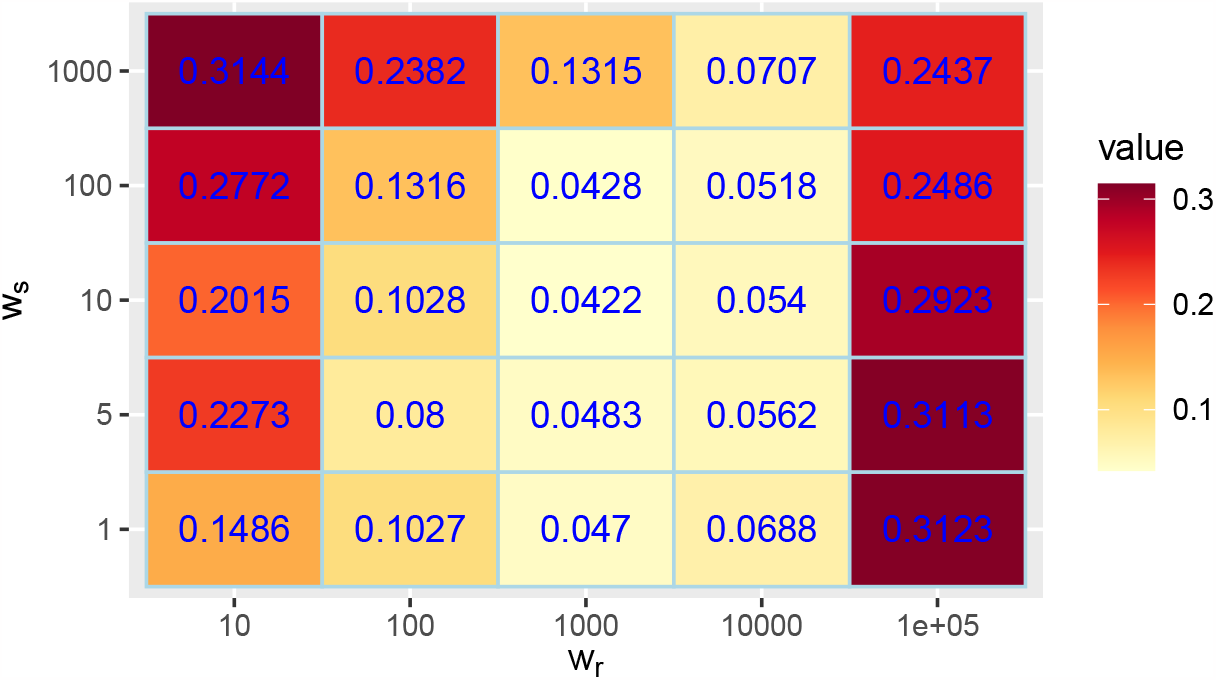
*RMSE* between the true *r*(*t*) and the one obtained by our method divided by the mean of the true *r*(*t*), using different choices for the regularization weights *w*_*r*_ and *w*_*s*_.

**Fig 14.**
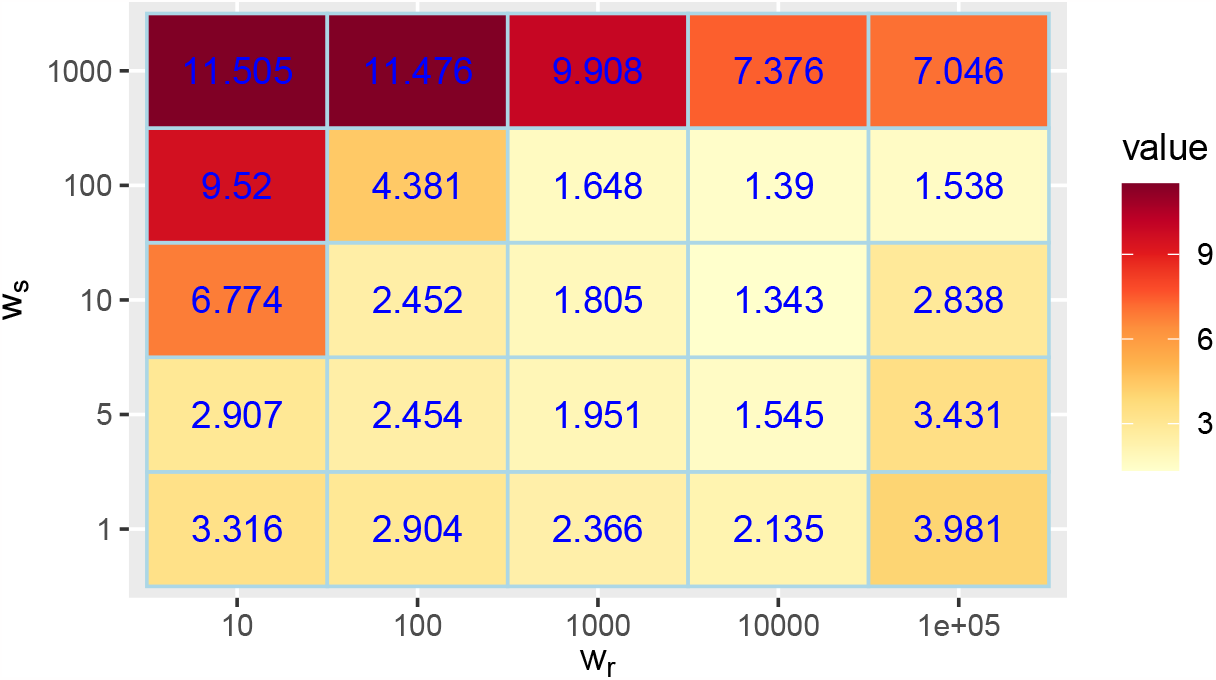
*RMSE* between the true *s*(*t*) and the one obtained by our method using different choices for the regularization weights *w*_*r*_ and *w*_*s*_.

## Conclusion

The results of the method are informative and realistic: we found that the proposed method enables a comparison between related epidemiological time series. A very good agreement of *r*(*t*)*f* (*t*) and *g*(*t* + *s*(*t*)) was generally observed in our experiments, for smooth *r*(*t*) and *s*(*t*), and the intervals where some disagreement was observed were explainable by real-world events. Our method also leads to propose generalizations of the renewal equation that enable relating curves of different data, thus generalizing the concept of reproducing number *R*_*t*_ to heterogeneous classes.

The main objection that can be raised by the method is its over-determination. Indeed, there are multiple solutions of (1), some trivial like *s*(*t*) = 0 and 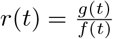, and many more. Indeed we can fix an arbitrary and smooth shift *s*(*t*), and obtain again a solution pair *r*(*t*), *s*(*t*) to (1) as 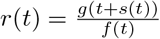. Thus, the joint imposition of regularity to *r*(*t*) and *s*(*t*) is crucial for our purposes. The good results obtained in the simulated study where a realistic ground truth for *r*(*t*), *s*(*t*) is properly recovered by the method, and the symmetry and transitivity of the method confirm that a sufficient regularity imposition on *r*(*t*) and *s*(*t*) properly solves the over-determination problem. From the modeling viewpoint, we verified that the behavior of *r*(*t*) and *s*(*t*) was interpretable and coherent with related results obtained by previous works.

## Data Availability

The data used are available at the Our World in Data web site

https://github.com/owid/covid-19-data/tree/master/public/data

